# Perceived concern and risk of getting infected with monkeypox among MSM: Evidence and perspectives from the Netherlands, July 2022

**DOI:** 10.1101/2022.09.26.22280354

**Authors:** Haoyi Wang, Kennedy J.I. d’Abreu de Paulo, Thomas Gültzow, Hanne M.L. Zimmermann, Kai. J. Jonas

## Abstract

The current monkeypox epidemic is most prevalent among men-who-have-sex-with-men (MSM). PrEP users and MSM with HIV (MSMHIV) are considered having the highest risk for monkeypox infection in the Netherlands and being targeted for monkeypox vaccination. Next to the epidemiological evidence, perceived concern/risk are also important in decision making about health behaviour uptake, e.g., vaccination uptake. It is thus relevant to examine which subpopulations among MSM consider themselves most at risk and most concerned about monkeypox. This study aimed to investigate this to complement and to help determining if the current measures to curb the epidemic are successfully targeted or not in the Netherlands. We conducted an online survey among 394 Dutch MSM. We first calculated the prevalence and standardised prevalence ratio (SPR) of high perceived concern/risk of monkeypox by the PrEP-use and HIV status. We then conducted two multivariable logistic regression analyses to investigate the perceived concern/risk of monkeypox and their potential socio-demographic/behavioural/health/psycho-social determinants. Among the included MSM, 52% showed high perceived concern and 30% showed high perceived risk of monkeypox. PrEP users (SPR=0.83) showed a significantly lower chance of perceived concern; and MSMHIV (SPR=2.09) were found to have a significant higher chance of perceiving high risk of monkeypox. In the multivariable logistic analyses, non-PrEP users (aOR=2.55) were more likely to perceive high concern, while MSM who were retired (aOR=0.23) and who had chemsex recently (aOR=0.63) were less likely to perceive high concern. MSMHIV (aOR=4.29) and MSM who had an unknown/undisclosed HIV status (aOR=6.07), who had attended private sex parties (aOR=2.10), and who knew people who have/had monkeypox (aOR=2.10) were more likely to perceive high risk of monkeypox. We found that a higher perceived risk (aOR=2.97) and a higher concern (aOR=3.13) of monkeypox were correlated with each other, more results see Table 2. In sum, only one-third of Dutch MSM considered themselves at a high risk of a monkeypox infection, and only half of them showed a high concern. We identified a potential discrepancy between the “actual risk” and the perceived risk and concern of monkeypox among MSM in this early stage of the monkeypox epidemic in the Netherlands, especially among PrEP users and MSMHIV. More refined public health communication strategies may be needed to improve the understanding and knowledge of the “actual risk” of monkeypox infections among these MSM sub-populations to encourage and facilitate an improved health behaviour uptake.

## Introduction

Monkeypox is a zoonotic disease described in the literature as a less-lethal relative of smallpox disease [1, 2], which is caused by the orthopoxvirus. On average, a few thousand cases occur in Africa on a yearly basis. However, the rapid development of its spreading outside the core area has put scientists and the public on high alert for monkeypox [2], and led to the declaration of a public health emergency by the World Health Organization (WHO) in July 2022 [3].

In the Netherlands, 1,166 cases were reported (assessed on August 31, 2022) with a majority occurring in Amsterdam [4]. Currently, the major infection routes were skin-to-skin contact and sexual contact facilitated by frequently changing partners [2, 5]. Most of the recent infections involved men who have sex with men (MSM) [4, 6]. MSM are thus considered to be the population with the highest risk of monkeypox [1]. Furthermore, the current monkeypox epidemic is quite likely to be stigmatised as another “gay epidemic” similar to the beginning of the human immunodeficiency virus (HIV) epidemic [5, 7]. This notion is being corroborated, in the Netherlands but also globally, due to some MSM sub-populations, such as MSM who use HIV pre-exposure prophylaxis (PrEP), and MSM with a diagnosed HIV being labelled as the most at risk population for monkeypox [8, 9], as evidence also shown that coinfection of monkeypox and HIV is possible [10]. Next to this epidemiological perspective, it is relevant to understand how members of affected communities perceive themselves in terms of monkeypox risk, and to examine which subpopulation among MSM considers themselves most at risk and most concerned about monkeypox. Such knowledge can complement both the epidemiological as well as the healthcare provider perspective and can help to determine if the current measures to curb the epidemic are sufficiently targeted or not. To close this gap, we investigated concern of and risk for a monkeypox infection among MSM in the Netherlands during the onset of the epidemic.

In psychosocial theories used to explain preventive behaviours towards infectious diseases, beliefs regarding risks and concerns are often linked to the use of such preventive behaviours. For example, in the Health Belief Model [11], perceived severity and perceived risk are two of the key determinants underlying the uptake of health behaviour, and have been associated with vaccination uptake, such as HPV and COVID-19, in previous studies [12-14]. However, for MSM, monkeypox is a novel health risk [15, 16], and individuals cannot easily fall back on previous knowledge to determine their infection risk, or to gauge how concerned they should be – as they can for other infections that have a longer history in this demographic [17, 18]. That said, both feelings of risk and concern might potentially determine MSM’s motivations to engage in protective behaviours, such as vaccination or risk reduction behaviours (e.g., reducing one’s number of sex partners) – both have been recommended for MSM [16, 19]. Two scenarios are possible: there could be 1) MSM who perceive themselves to be at risk of monkeypox who are actually at risk and should therefore be concerned; 2) MSM who do *not* perceive themselves to be at risk, while they are actually at risk and thus pay less attention to the topic, which may lead to more infections [20]. Due to a lack of knowledge in regards to MSM’s feelings of risk and concern, public health and health communication interventions may not be efficiently targeted which might make it more likely that the target group misses opportunities for monkeypox prevention. In the long run, this may result in protective interventions not being used by MSM sub-populations that are at the highest risk. It is thus important to understand how MSM understand this epidemic and perceive themselves in terms of concerns and risks in relation to monkeypox, especially when they are considered as the most-at-risk population. Consequently, more appropriate and efficient public health interventions can be designed and implemented to prevent transmissions among MSM.

Therefore, in addition to unravelling the epidemiologic profile of the perceived concern and risk of monkeypox among MSM, it is also important to identify which MSM sub-population are more likely to perceive concern and risk of monkeypox, together with their socio-demographic, behavioural/health, and psycho-social profiles, to understand whether there is a match of perceived concern and risk and actual risk determinants. A previous study has provided insights into the determinants of monkeypox vaccination intention among MSM in the Netherlands [16]. Even though perceived concern and risk were associated with vaccination intention [16], it is not appropriate to assume the reported determinants also play similar roles in terms of the perceived concern and risk for monkeypox, given the different natures of these endpoints.

Therefore, this study aimed to investigate the perceived concern and risk of monkeypox among MSM, and to investigate determinants of perceived concern and risk for monkeypox in MSM in the Netherlands to better understand the appraisal of the monkeypox epidemic among this population at the early stages of the current monkeypox epidemic.

## Materials and Methods

### Study design and participants

This study has a cross-sectional design. We conducted an online survey among a convenience sample consisting of 394 MSM in July 2022, of which 257 were from a cohort established in 2017 [21] and 137 MSM from an online gay dating app. In this study, we only included data from respondents who indicated they were living in the Netherlands. This study was assessed and approved by the Ethics Review Committee Psychology and Neuroscience of Maastricht University (ref.188_11_02_2018_S32). Informed consent was provided by all participants. For more information on the design of the online survey, please see our previous study which used the same data for other endpoints [16].

### Measures

#### Outcomes measures

All measures were self-reported. Participants were asked they 1) “How worried are you to catch monkeypox yourself” (hereinafter perceived concern), and 2) “How likely is it that you will catch monkeypox yourself” (hereinafter perceived risk). These two endpoints were measured using an 1-5 Likert scale (with 1 = “Very low” and 5 = “Very high”).

Following our previous study on monkeypox vaccination intention and self-isolation intention [16], we also assessed socio-demographic, behavioural/health, and psycho-social determinants.

#### Socio-demographic determinants measures

For socio-demographic determinants, age in this study was dichotomised as “younger than 45-year-old” and “older than 45-year-old”, given the median age group in our participants was 45-55 year-old (for more information, see our previous study [16]). Relationship status was categorised into “Single”, “Single but dating”, “In a monogamous relationship”, and “In an open/polygamous relationship”. Education was categorised into “Lower than Bachelor”, “Bachelor”, “Master”, and “PhD or higher”. Employment status was categorised into “Employed”, “Unemployed or receiving social welfare”, “Retired”, and “Student”. Migration status was categorised into “No migration status”, “First generation immigrant”, and “Second generation immigrant”. For place of residence, given the fact that most of the current monkeypox cases were diagnosed in Amsterdam and surrounding regions, we categorised this variable into “The main urban area of the Netherlands” and “The rest of the country”. The main urban area of the Netherlands entails the agglomeration of cities in the west of the Netherlands, in particular Amsterdam, Utrecht, Leiden, The Hague, and Rotterdam (in Dutch: Randstad). As the economic and political centre of the Netherlands, the Randstad accounts for approximately 50% of the national population [22].

#### Behavioural/health determinants measures

For behavioural/health determinants, number of sex partners in the previous six months were categorised into “None”, “1”, “2 to 6”, “7 to 15”, and “More than 15”. HIV status was measured based on the HIV diagnosis of the participants, and categorised into “Negative”, “having a positive diagnosis”, and “Unknown or not disclosed”. PrEP use status was categorised into “current PrEP users” and “PrEP naïve or PrEP discontinued” which indicates a non-PrEP using status.

We also measured past behaviours in the previous 6 months which may associated with a higher risk of monkeypox infection such as substance use status and gay subculture/sexual activities [23]. For substances use, we measured whether participants ever used any type of substance in the previous 6 months (“Ever” / “Never”); ever used any recreational drug, such as THC, MDMA, ecstasy etc., in the previous 6 months (“Ever” / “Never”); ever had chemsex, such as using crystal meth/tina, GHB, ketamine etc. in the previous 6 months (“Ever” / “Never”); ever used poppers in the previous 6 months (“Ever” / “Never”); ever used erectile dysfunction drugs, such as Viagra® or Kamagra®, in the previous 6 months (“Ever” / “Never”); ever used alcohol in the previous 6 months (“Ever” / “Never”). For gay subculture/sexual activities, we measured whether the participants ever visited a gay sauna in the previous 6 months (“Ever” / “Never”); ever visited a darkroom in the previous 6 months (“Ever” / “Never”); ever visited a circuit party in the previous 6 months (“Ever” / “Never”); ever visited a Pride event in the previous 6 months (“Ever” / “Never”); ever visited a gay dance club in the previous 6 months (“Ever” / “Never”); ever attended to a private sex party in the previous 6 months (“Ever” / “Never”); and ever visit a fetish event in the previous 6 months (“Ever” / “Never”).

#### Psycho-social determinants measures

For psycho-social determinants, we measured whether the participants knew anybody who has/had monkeypox (“Yes” / “No”). We also measured their perceived problematic consequences of monkeypox: “how problematic is it to catch monkeypox” using an 1-5 Likert scale (with 1 = Not problematic at all” and 5 = “Very problematic”). To investigate the association and potential correlation between the two endpoints, we included the perceived risk and concern of monkeypox for the endpoint of perceived concern and the endpoint of perceived risk of monkeypox respectively using the -5 Likert scale (with 1 = “Very low” and 5 = “Very high”).

### Statistical analysis

Given the relatively very small proportion of the participants showing both very high perceived concern and risk of monkeypox in this study (more details see the Results and Figure 1) and to better inform public health actions, following the analysis strategy from our previous studies [16, 19], we dichotomised the two outcome endpoints as “High/very high (scale 4 and 5)” and “The rest of the scale points (scale 1-3)” for modelling analysis.

**Figure 1.**
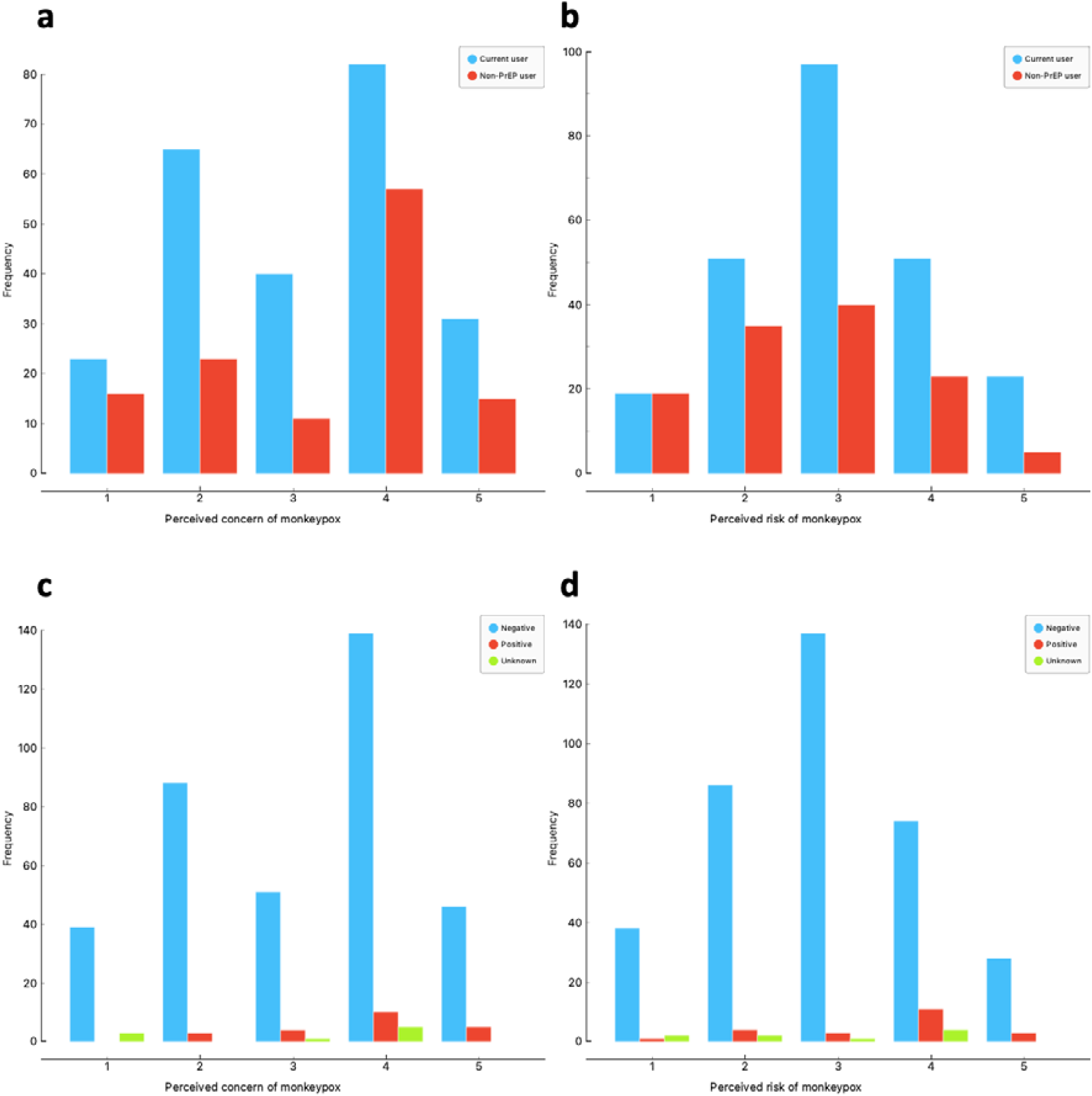
Distribution of a) monkeypox perceived concern by PrEP use status, b) monkeypox perceived risk among MSM by PrEP use status, c) monkeypox perceived concern by HIV status and d) monkeypox perceived risk among MSM by HIV status. Note: two endpoints were measured on a 1-5 Likert scale (with 1 = “Very low” and 5 “Very high”)

#### Descriptive analysis

We first estimated and compared the crude prevalence and standardised prevalence ratio (SPR) of the perceived concern and risk of monkeypox by PrEP use status and HIV status, given that MSM who use PrEP and who are living with HIV are the ones of the current monkeypox vaccination priority populations in the Netherlands [8]. SPR allows us to compare the risk levels in different sub-populations if one sub-population has higher (SPR > 1), equal (SPR = 1) or lower (SPR < 1) probability than the overall prevalence in the total study population [22].

#### Multivariable logistic regression modelling

We then conducted two multivariable logistic regressions with socio-demographic, behavioural/health, and psychosocial determinants for each endpoint. Potential collinearity was not found, for the analysis see [16]. Firstly, we conducted an univariable logistic regression with each included determinant. All determinants with a p<0.10 identified in the univariable modelling analyses were retained in the multivariable model, given the relatively small sample size. All determinants with a p<0.05 in the multivariable model were considered statistically significant. All analyses were conducted in R (version R 4.2.1) (R Foundation for Statistical Computing, Vienna, Austria).

## Results

### Study population characteristics

Of the included 394 MSM, 43% were below the age of 45, 61% were living in Randstad of the Netherlands, 6% were living with HIV, 66% were currently using PrEP, 26% had chemsex and 40% had attended to private sex parties in the previous six months, and 17% knew people who have/had monkeypox (see Table 1 for other study population characteristics).

**Table 1.**
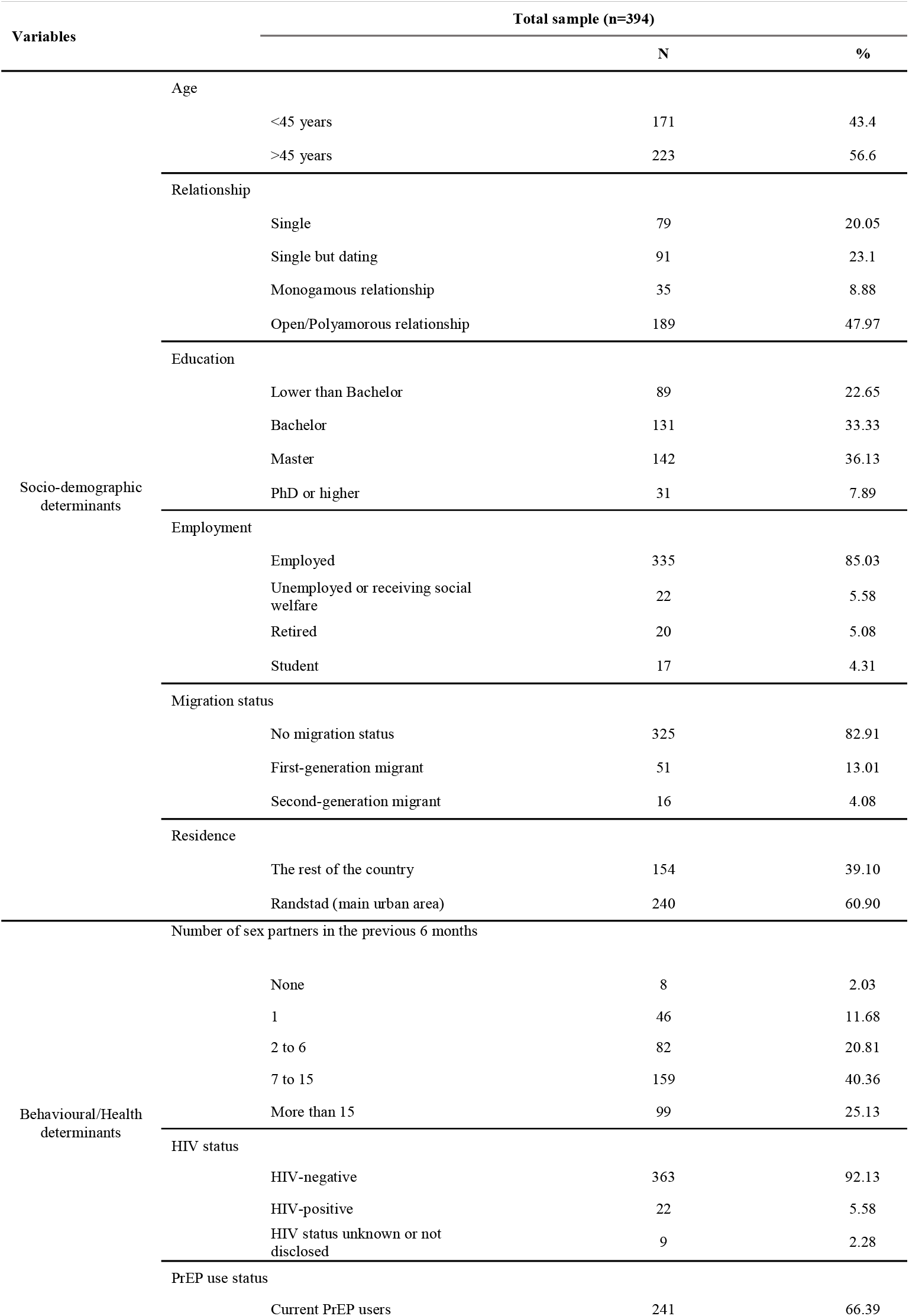
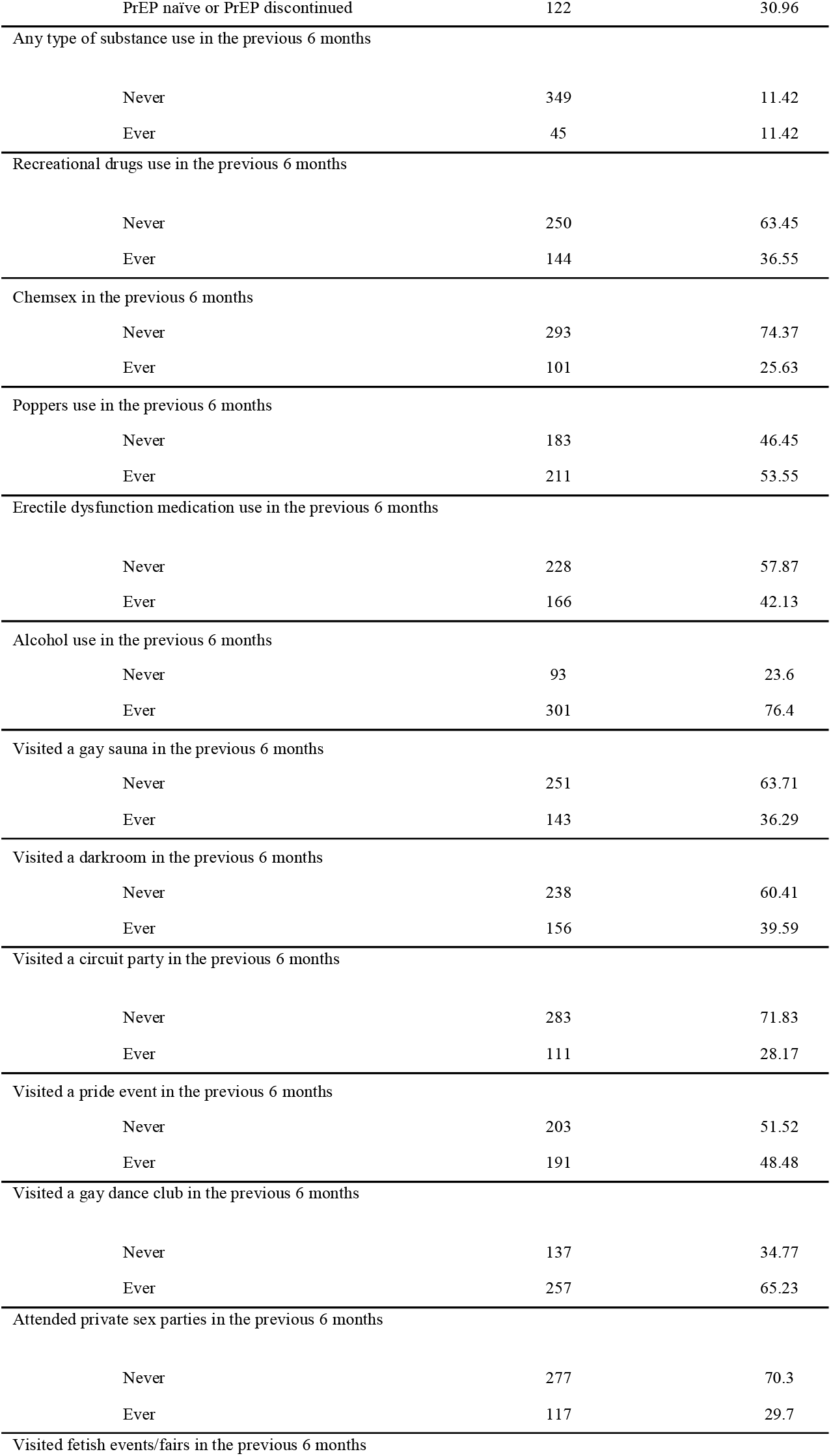
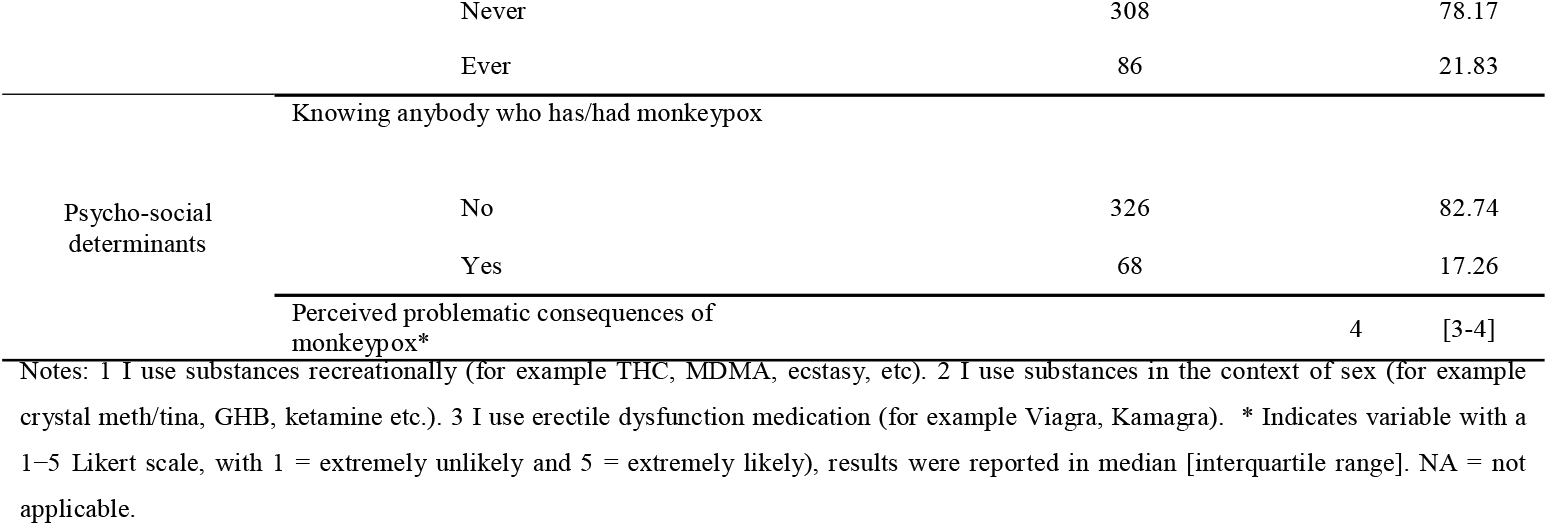
Study characteristics.

### Perceived concern and risk of monkeypox among MSM

Among the included MSM, 39% showed “high” and 13% “very high” perceived concern, and 23% showed “high” and 8% “very high” perceived risk. Figure 1 summarised the frequencies of the observation of a) monkeypox perceived concern by PrEP use status, b) perceived risk among MSM by PrEP use status, c) monkeypox perceived concern by HIV status, d) perceived risk among MSM by HIV status.

After combining the “High” and “Very high” scales for the endpoints, among the total sample, 52% showed high/very high perceived concern, and 30% showed high/very high perceived risk of monkeypox. When comparing results by PrEP use status, only current PrEP users (prevalence=47%, SPR=0.83) showed a significantly lower probability of perceived concern of monkeypox compared to non-PrEP users. No significant difference of probability of high/very high perceived risk of monkeypox was found between current PrEP users (prevalence=31%, SPR=1.09) and non-PrEP users (prevalence=23%, SPR=0.82). When comparing results by HIV status, no significant difference of probability of high/very high perceived concern of monkeypox was found between MSM living without HIV (prevalence=51%, SPR=0.98), MSM living with HIV (prevalence=68%, SPR=1.31), and MSM whose HIV status were unknown or not disclosed (prevalence=56%, SPR=1.06). Only MSM living with HIV (prevalence=64%, SPR=2.09) were found to have a significant higher probability of perceived high/very high risk of monkeypox compared to MSM with other HIV status. See Table 2 for more information of the estimated prevalence and SPR of perceived concern and risk by PrEP use status and HIV status.

**Table 2.** Prevalence and standardised prevalence ratio of perceived concern and risk of monkeypox among MSM in the Netherlands, July 2022.

| Sub-population | Perceived concern of monkeypox<br>(High/very high vs. rest of scale) * |  |  |  |  | Perceived risk of monkeypox<br>(High/very high vs. rest of scale) * |  |  |  |  |
| --- | --- | --- | --- | --- | --- | --- | --- | --- | --- | --- |
|  | n | Prevalence (%) | 95% CI | SPR | 95% CI | n | Prevalence (%) | 95% CI | SPR | 95% CI |
| **Total sample (N=394)** | 205 | 52.03 | 47.10;56.92 | NA | NA | 120 | 30.46 | 26.12;35.17 | NA | NA |
| **PrEP users (N=241)** | 113 | 46.89 | 40.69;53.19 | 0.83 | 0.68;0.99 | 74 | 30.71 | 25.22;36.79 | 1.09 | 0.86;1.36 |
| **Non-PrEP users (N=122)** | 72 | 59.02 | 50.14;67.34 | 1.05 | 0.82;1.30 | 28 | 22.95 | 16.38;21.16 | 0.82 | 0.54;1.15 |
| **HIV positive (N=22)** | 15 | 68.18 | 47.31;83.63 | 1.31 | 0.73;2.05 | 14 | 63.64 | 42.95;80.27 | 2.09 | 1.14;3.32 |
| **HIV negative (N=363)** | 185 | 50.96 | 45.84;56.07 | 0.98 | 0.84;1.13 | 102 | 28.10 | 23.72;32.93 | 0.92 | 0.75;1.11 |
| **HIV status unknown/undisclosed (N=9)** | 5 | 55.56 | 26.67;81.12 | 1.06 | 0.34;2.21 | 4 | 44.44 | 18.88;73.33 | 1.45 | 0.38;3.23 |

Note: CI: confidence interval; NA: not applicable; PrEP: pre-exposure prophylaxis; SPR: standardised prevalence ratio. \* 1 □ – □ 5 Likert scale, with 1 □ = □ extremely unlikely and 5 □ = □ extremely likely)

### Determinants of perceived concern and risk among MSM

For perceived concern, no socio-demographic determinants were found to be statistically associated with the end point. Among behavioural/health determinants, MSM who did not use PrEP (adjusted odds ratio (aOR)=2.55) were more likely to show high/very high levels of concern about a possible monkeypox infection. On the other hand, MSM who had chemsex in the previous six months (aOR=0.44) were less likely to show a high/very high concern about a possible monkeypox infection. Among psycho-social determinants, MSM who had a higher perceived risk of monkeypox (aOR=3.26) were more likely to perceive more concern of getting infected of monkeypox.

For perceived risk, similarly, no socio-demographic determinants were found to be associated with this end point. Among behavioural/health determinants, MSM who had a diagnosed-HIV status (aOR=4.29) and unknown-HIV status (aOR=6.07), and who had ever attended private sex parties in the previous 6 months (aOR=2.10) perceived themselves at higher risk for monkeypox. PrEP use status was not associated with perceived risk univariably and multivariably. Among psychosocial determinants, MSM who knew anybody who had monkeypox (aOR=2.60) and who perceived a higher concern of monkeypox (aOR=3.24) perceived themselves at higher risk for monkeypox. For results obtained from univariable logistic regression analyses for both endpoints, see Table 3.

**Table 2.**
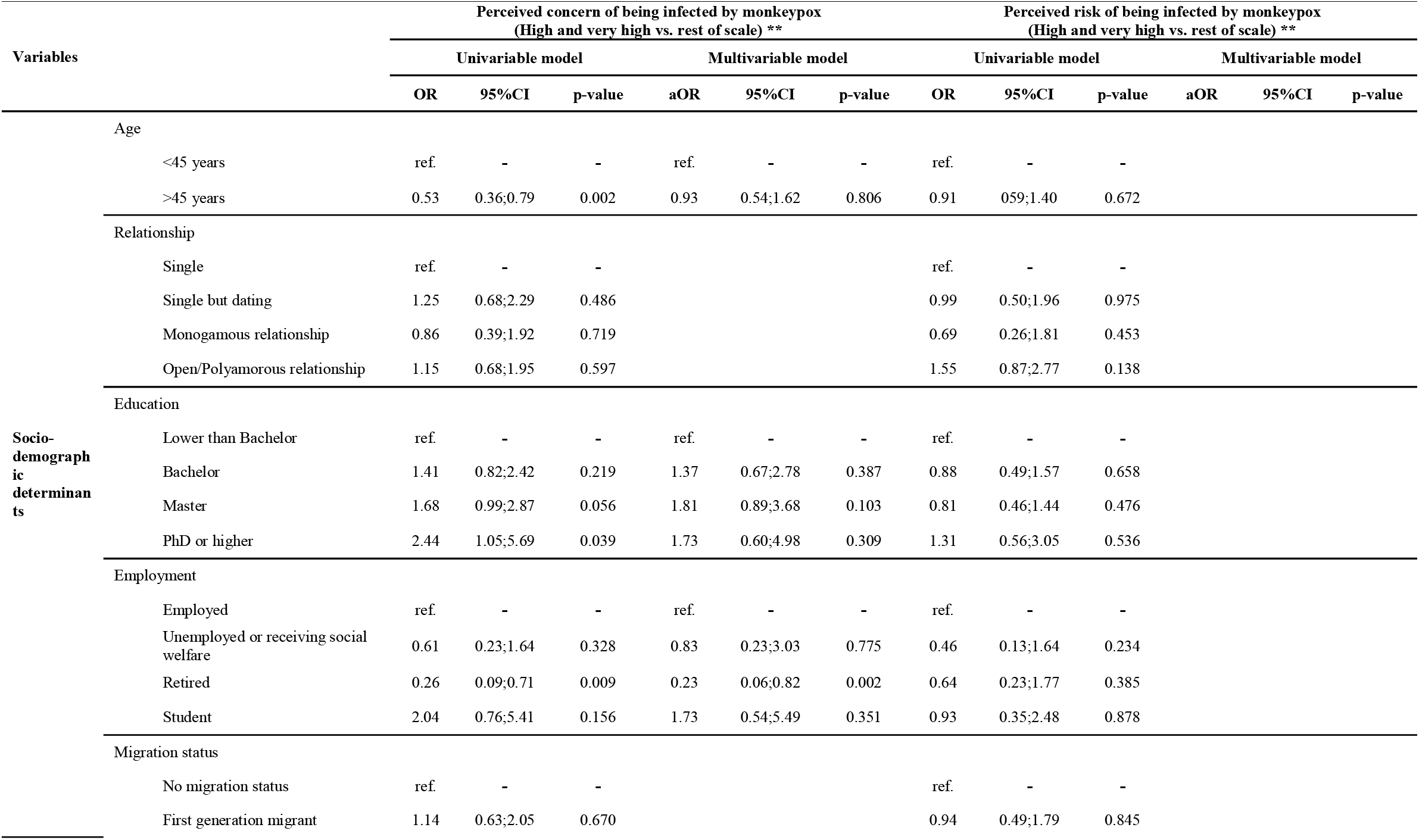
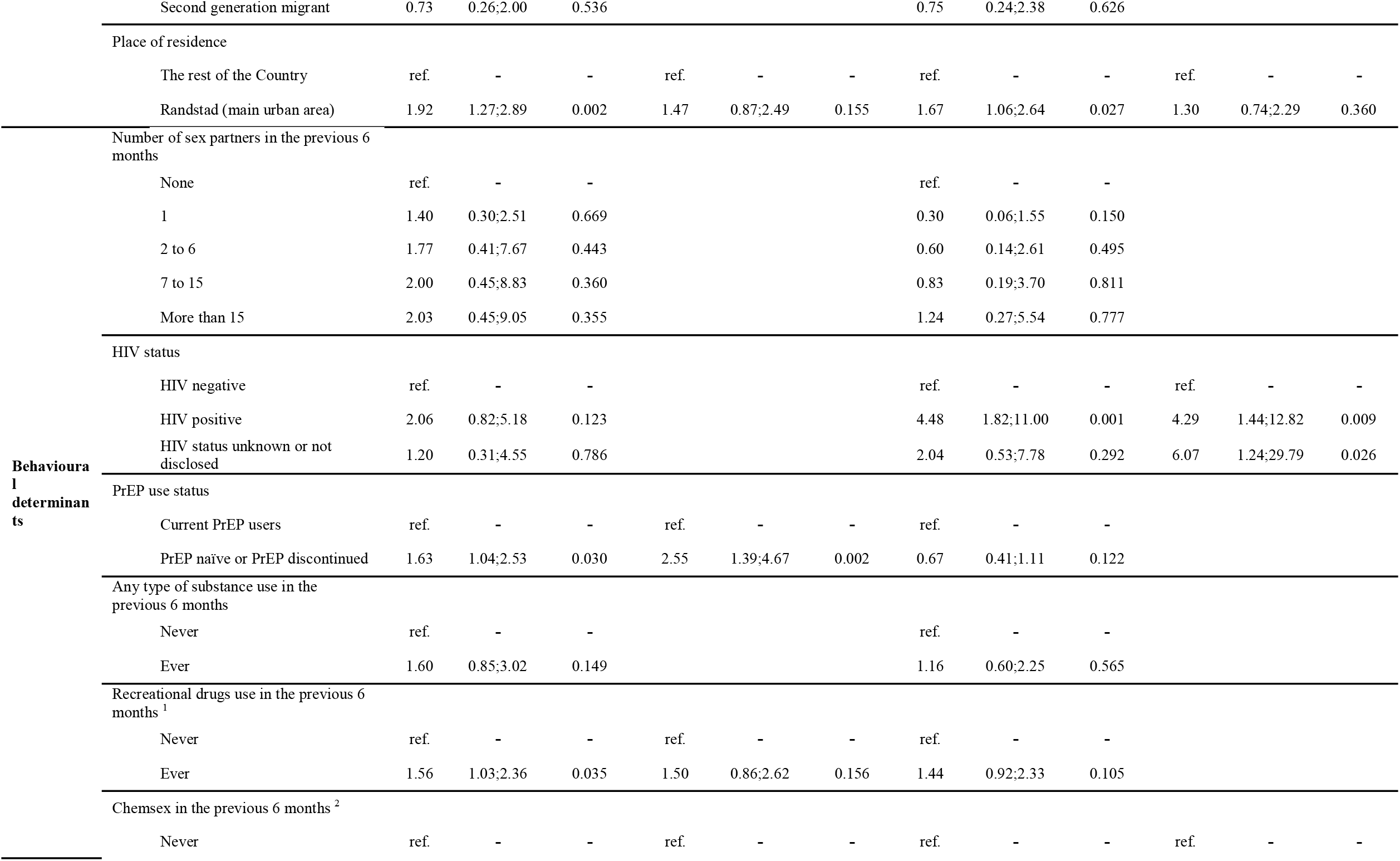
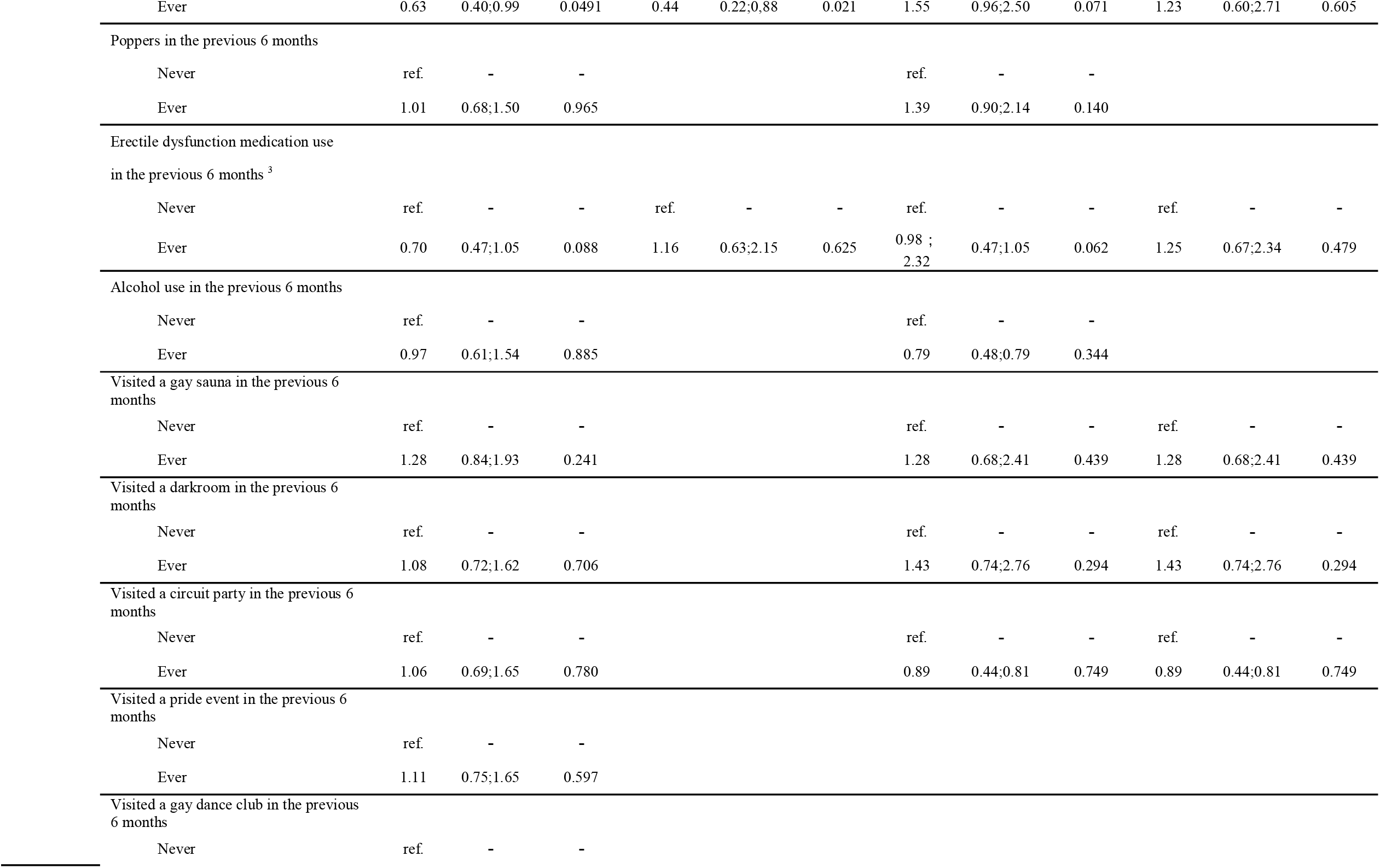
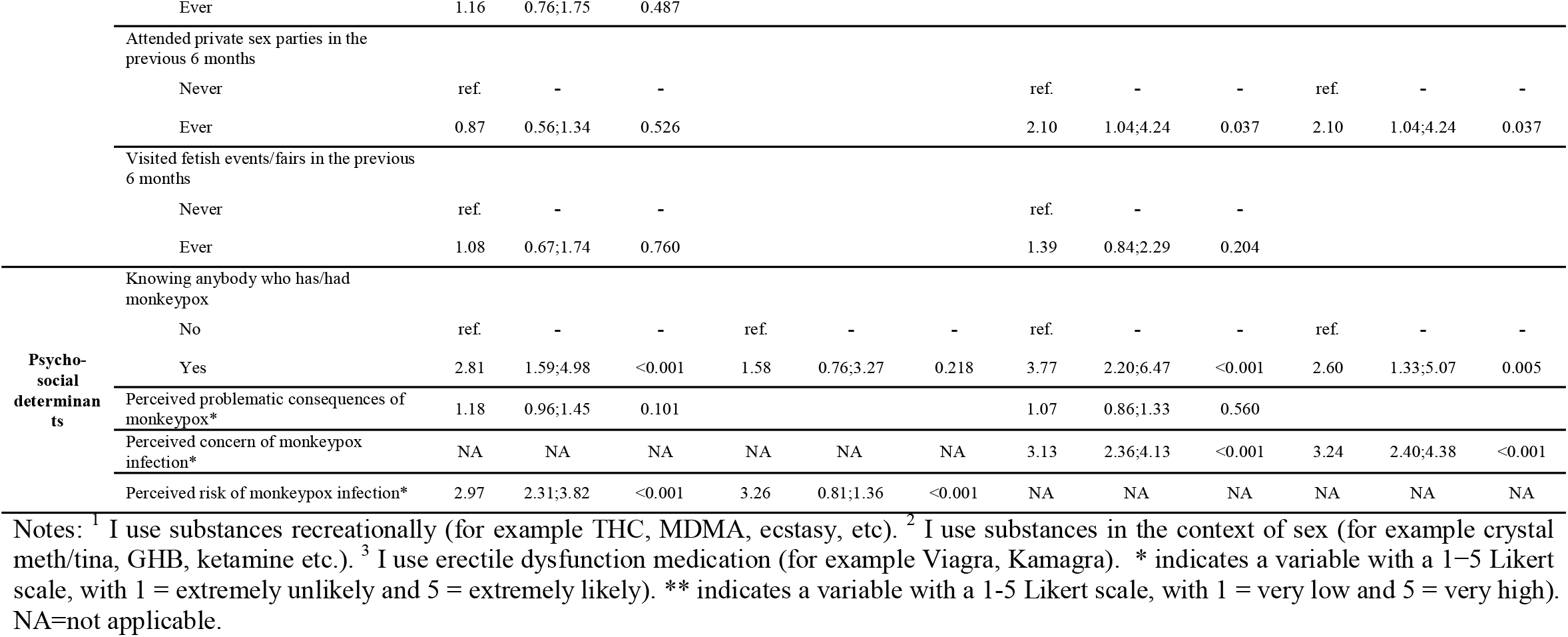
Determinants of perceived concern and perceived risk of getting monkeypox among MSM in the Netherlands, July 2022.

## Discussion

To our knowledge, this study is the first study to report perceived concern and risk of monkeypox and their determinants among MSM. Our findings based on 394 MSM in the Netherlands showed that 52% of our respondents showed high/very high levels of perceived concern about monkeypox, but only 30% perceived themselves to be high/very high risk for monkeypox, based on data collected at an early stage of monkeypox epidemic prior to vaccination implementation. In addition, these results also highlight there may be a discrepancy between the “actual risk” according to an epidemiolocal viewpoint [1, 5, 15, 24, 25] and perceived risk and concern among the population at risk among MSM living in the Netherlands. Also, there seems to be a mismatch between MSM’s level of concern and level of risk. Thus these results indicate a lack of understanding and knowledge of the ongoing monkeypox epidemic among MSM in the Netherlands.

Not surprisingly, some of the included psycho-social determinants play a role for both endpoints in our analysis such as knowing people with monkeypox. Proximity to the health threat is a typical determinant for concern and risk perception [26]. Our results also indicated that perceived concern in relation to monkeypox and perceived risk of monkeypox among MSM were associated with each other positively, similar to perceived severity and perceived susceptibility in the Health Belief Model [11], which could indicate that beliefs in relation to concerns and risks about monkeypox should be targeted and included simultaneously. Another potential explanation for this finding could be that two endpoints do in fact reflect an underlying psychological construct, such as perceived threat in the Health Belief Model. However, we also identified other determinants which may influence both determinants among MSM differently.

While non-PrEP users and current PrEP users showed similar levels of perceived risk of Monkeypox, non-PrEP users showed a significant higher concern of being infected with monkeypox compared to current PrEP users. This may indicate a potential interaction effect from the current monkeypox vaccination strategy which is predominantly focusing on PrEP using MSM [8]. As non-PrEP users cannot access the monkeypox vaccine at the moment in the Netherlands, they may consider themselves not protected against monkeypox epidemic, which may thus lead to a higher perceived concern of monkeypox infection even when sharing similar risk beliefs as current PrEP users. Another reason may be the lack of knowledge and understanding of monkeypox among MSM, especially among the current PrEP users. After being free of risk of acquiring HIV by using PrEP [27], PrEP users may not consider themselves to still be at risk for monkeypox. This finding dovetails nicely with other results showing that risks to acquire other STIs while using PrEP are discounted, too [18]. Given the significantly lower perceived concern of monkeypox infection among PrEP users, more public health communication may be needed to improve the current understanding of monkeypox as a novel health risk, which cannot be covered by PrEP. However, before this, further research should explore the underlying reasons for the differences in perceived risks.

Similarly, but in an opposite direction, HIV status also played differential roles in the level of perceived concern and risk of monkeypox among MSM. A positive and unknown/non-disclosed HIV status predicted a higher likelihood of perceived risk but not a higher perceived concern of monkeypox among MSM. This finding could reflect the awareness for risky sexual behaviour that a higher risk of monkeypox infection is perceived among this sub-population of MSM, however, given their experience of HIV infection, they may not regard infecting with monkeypox is as problematic as HIV, similar to the perception of being infected with other STIs among people living with HIV [28]. In addition, another reason for this finding may be the unknown risk and pathways of developing a more severe disease outcomes among people living with HIV [25]. Even though a higher risk of monkeypox infection is perceived among this population, it is still currently unknown whether an HIV infection alters a person’s risk of acquiring monkeypox after exposure [25], MSM with HIV or unknown HIV status may not develop a higher perceived concern for this infection.

While our previous study reported no behavioural determinants to be associated with monkeypox vaccination intention among MSM in the Netherlands [16], we identified several past behaviours to be associated with the perceived concern and risk of monkeypox among MSM both univariably and multivariably, indicating that past behaviours are associated with vaccination intentions and perceived concern and risk differently. For example, our study showed that people who had chemsex in the previous six months showed a lower likelihood of perceiving a high/very high concern of monkeypox. One possible reason maybe that MSM who engage in chemsex activity tend to underestimate their risk for infections that can be transmitted via sexual contacts in general [29]. Our study suggests a potential similar mechanism behind the lower perceived risk of monkeypox among MSM who had chemsex. In addition, based on the multivariable model, not surprisingly, MSM who attended private sex parties in the previous six months were more likely to perceived themselves having high/very high risk for monkeypox. An elevated number of sex partners and sexual activities may be associated with this behaviour (a potential negative association was shown between a higher number of sex partners and never attend private party were shown in our collinearity analysis [16]), and thus lead to a higher perceived risk of monkeypox, which indicates that for this sub-population of MSM, they indeed perceived a higher risk when they are actually at risk for monkeypox. Therefore, there is at least some evidence that certain sub-populations gauge their risk correctly.

Although MSMs’ place of residence was found to be associated with both Monkeypox perceived concern and risk univariably, such effects disappeared after adjusting for other determinants. Given the fact that most of monkeypox cases were reported from Amsterdam [4], we hypothesised a higher perceived concern and risk among MSM from the Randstad. However, no differences in both endpoints between Randstad and the rest of the country were found. One reason may be due to the ecological fallacy. Even though Amsterdam rests within the Randstad region, the internal heterogeneity of the likelihood of perceived concern and risk between different cities within Randstad can be masked when aggregating data on the Randstad level. It could also be the MSM from the Randstad differ regarding the other included variables compared to MSM from other regions which led to the effects disappearing in the multivariable models.

### Limitations and recommendations for future research

Despite the novelty and timely communication of this study on the perceived concern and risk of monkeypox among MSM with evidence and perspectives from the Netherlands, there are a few limitations that are applicable to our study.

First, we suggested a potential interaction between vaccination strategy and PrEP use status. However, due to insufficient data, future studies are warranted and should therefore further investigate such an interaction with both more qualitative and empirical evidence. Second, our data were collected at the beginning stage of the monkeypox epidemic while the comprehensive public health communication and public health measures (i.e., vaccination) were just starting up. This may have influenced MSMs’ knowledge of monkeypox, and eventually led to underestimated levels of perceived concern and risk of monkeypox among MSM. An updated assessment at a later stage could be warranted to measure the change in perceived concern and risk of monkeypox since more information has been shared with the public in the meantime. Although, the future case development can be taken into account. Third, given the relatively small sample size in this study, the power of our models may be limited. For example, our models did not have the power to investigate the covariations from the geo-location on a finer geographic scale such as public health services regional level (25 in total), or municipality level (345 in total). Our results on the spatial perspective may thus be limited and not comprehensive to support local monkeypox prevention. Therefore, future studies could zoom in on a finer geographical level, to provide more concise estimations with a larger sample size. Lastly, given that our data were self-reported, our data were thus not devoid of information biases, especially on the past risky behaviours. This may result in biased parameters.

## Conclusions

In conclusion, only a small proportion of MSM living in the Netherlands considered themselves at a high/very high risk of monkeypox infection, and around half of them showed a high/very high concern for it. We found that the current PrEP users and non-PrEP users shared similar perceived risk of monkeypox infection, but non-PrEP users were more concerned about the monkeypox infections. A potential discrepancy between the “actual risk” and the perceived risk and concern of monkeypox, may exist among MSM in this early stage of the monkeypox epidemic. Public health professionals should therefor put more effort on improving the understanding and knowledge of the “actual risk” of monkeypox infections among these MSM sub-populations to encourage and facilitate an improved health behaviour uptake.

## Data Availability

Data are available upon request.

